# Clinical nomogram using novel CT based radiomics predicts survival in non-small cell lung cancer patients treated with SBRT

**DOI:** 10.1101/2022.06.21.22276718

**Authors:** Eashwar Somasundaram, Raoul R. Wadhwa, Adam Litzler, Rowan Barker-Clarke, Peng Qi, Gregory Videtic, Kevin Stephans, Nathan A. Pennell, Daniel Raymond, Kailin Yang, Michael W. Kattan, Jacob G. Scott

## Abstract

**Introduction:** Improved survival prediction and risk stratification in non-small cell lung cancer (NSCLC) would lead to better prognosis counseling, adjuvant therapy selection, and clinical trial design. We propose the PHOM (persistent homology) score, the radiomic quantification of solid tumor topology, as a solution.

**Methods:** Patients diagnosed with stage I or II NSCLC primarily treated with stereotactic body radiation therapy (SBRT) were selected (*n* = 554). The PHOM score was calculated on each patient’s pre-treatment CT scan (10/2008 to 11/2019). PHOM score, age, sex, stage, Karnofsky Performance Status (KPS), Charlson-Comorbidity Index (CCI), and post-SBRT chemotherapy were predictors in the Cox proportional hazards models for overall and cancer-specific survival. Patients were split into high and low PHOM score groups compared using Kaplan-Meier curves for overall survival and cumulative incidence curves for cause specific death. Finally, we generated a validated nomogram to predict overall survival, publicly available at https://eashwarsoma.shinyapps.io/LungCancerTDATest/.

**Results:** PHOM score was a significant predictor for overall survival (HR: 1.17, 95% CI: 1.07−1.28) and was the only significant predictor for cancer-specific survival (1.31, 95% CI: 1.11−1.56) in the multivariable Cox model. The median survival for the high PHOM group was 29.2 months (95% CI: 23.6−34.3), which was significantly worse compared to the low PHOM group (45.4 months, 95% CI: 40.1−51.8, *p <* 0.001). The high PHOM group had a significantly greater chance of cancer-specific death at post treatment month 65 (0.244, 95%CI: 0.192−0.296) compared to the low PHOM group (0.171, 95% CI: 0.123−0.218, *p* = 0.029).

**Conclusions:** The PHOM score is associated with cancer-specific survival and predictive of overall survival. Our developed nomogram can be used to inform clinical prognosis and assist in making post-SBRT treatment considerations.

## 1 Introduction

Approximately 230,000 cases of lung cancer are diagnosed, and 135,000 patients die from lung cancer each year in the US^1^. Proportionally, 85% of all lung cancer cases are non-small cell lung cancer (NSCLC)^2^. Primary therapy has traditionally been surgical resection; however, stereotactic body radiation therapy (SBRT) is now an option for patients with inoperable tumors^3^. For all cancers, clinical prognostication is essential for therapy selection. This is especially important in lung cancer, for which 5-year survival is highly variable in NSCLC ranging from nearly 70% for patients with stage I disease down to 10% for patients with metastatic disease^4^. While early stage has an overall positive prognosis, approximately 30% of patients do not survive at 5 years. This indicates a need to develop better risk stratifying metrics.

Risk calculators are clinical decision support tools that quantify prognosis. They typically use histopathologic and clinical covariates to predict outcomes. Risk calculators incorporating molecular and clinical information have been developed for lung cancer^5^. While clinical data is easily obtained, molecular data often poses a challenge outside of tertiary medical centers. In contrast, imaging data is far more accessible. Radiomics–the science of extracting and analyzing features from imaging data–has been increasingly used to predict clinical outcomes in cancer^6^. While radiomics already offers a wide toolset for image analysis, we propose a valuable extension with persistent homology.

Persistent homology quantifies the global topology, or overall structure, of big data^7^. Its high level approach provides the benefit of discriminating signal from noise. Oncology already features structure analysis similar in spirit to persistent homology. Glandular shape analysis through Gleason scoring is used to grade prostate cancer severity^8^.

We hypothesize that malignant and benign tissue exhibit distinct topology on clinical imaging. Malignant tumors are typically diffusely spread, heterogeneous in composition, and may possess necrotic cavities. In contrast, benign tumors are well-circumscribed and likely to be homogeneous in appearance^9^. Persistent homology quantifies these observations to permit their use in predictive models. Of note, persistent homology has already been introduced to radiomics in the study of glioblastoma and liver cancer^10,11^.

Within lung cancer, persistent homology has shown promise in predictive modeling via a metric named the PHOM score^12^. In our previous study, we demonstrated that the PHOM score is associated with overall survival in patients with NSCLC treated with surgery or radiation. In this study, we utilize the advantage of using our institutional data to curate a cohort with greater resolution of clinical detail. We selected patients treated with SBRT since their tumor margins on imaging are contoured for treatment purposes. The primary aim of this study was to assess our hypothesize that the PHOM score can predict overall survival in patients with NSCLC treated with SBRT. The secondary aim was to create clinical risk groups using the PHOM score. Here, we validate the PHOM score using a large prospectively collected, IRB approved dataset from our institution including 554 patients treated with SBRT for primary lung cancers from 2008-2019. We combine our PHOM score with clinical covariates to develop a prognostic nomogram that predicts duration of overall survival post-SBRT. This prognostic tool would assist providers and their patients in making both personal and medical decisions. We also demonstrate the PHOM score’s ability to stratify patients into risk groups. Risk stratification would assist adjuvant therapy selection at the individual patient level and clinical trial design at a more global level.

## 2 Materials and Methods

### 2.1 Source of Data and Participants

We developed a data pipeline that calculated the PHOM score of computed tomography (CT) tumor scans to create validated survival regression models. Methodology to generate a PHOM score using tumor CT scans is described in our previous work^12^. Only patients with early stage (I or II) biopsy proven primary non-small cell lung cancer with no prior treatment history were eligible for this study. We utilized a single institution prospectively collected dataset of 803 patients receiving SBRT with pretreatment patient scans (scan dates ranging 10/2008 to 11/2019) with corresponding clinical data. Clinical data including dates of diagnosis, last followup, and treatment were obtained from Epic chart review. Patients had uniform free-breathing CT parameters with slice thickness of 3mm.

After excluding 249 patients with metastatic disease, nodal disease, additional primary tumors, and patients who received pretreatment, 554 patients met inclusion criteria (**Figure 1A**) All patients received SBRT as their primary treatment for local control.

**Figure 1.**
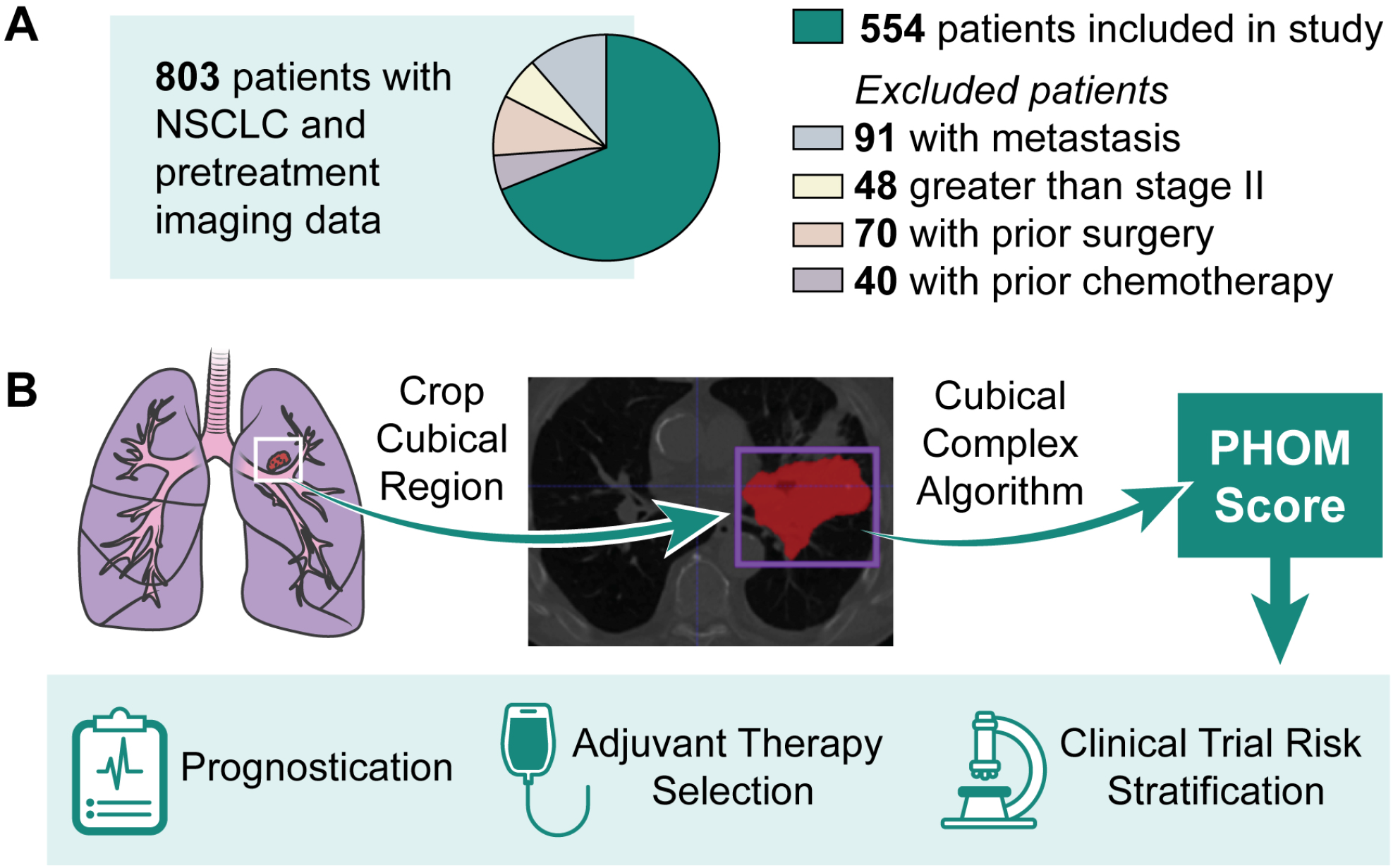
Conceptual Overview of Translating Utility of PHOM Score. **(A)** 803 patients were in our database with imaging and clinical data for NSCLC treated primarily with SBRT. 249 patients were excluded based on their stage or treatment history. **(B)** The PHOM Score is algorithmically calculated from a tumor segmented by a clinician. Note, this particular cross section demonstrates a large tumor greater than stage II but was selected for visualization purposes. Only stage II tumors and below were selected for analysis. **(C)** We envision the PHOM Score to have utility in prognostication, therapy selection, and risk stratification.

### 2.2 Model Development

Each patient’s primary lung tumor had been segmented by a radiation oncologist. For each CT scan, we used the segmentation coordinates to crop scan to only include tumor tissue. The PHOM score was calculated from this data object (**Figure 1B**). As previously described, this metric reflects the number of disconnected foci of pixels with similar grayscale values in the segmented tumor object^12^.

We demonstrate the PHOM score’s potential clinical utility through a Cox proportional hazards model and nomogram (**Figure 1C**). PHOM score, age, sex, Karnofsky Performance Status (KPS), Charlson Comorbidity Index (CCI), overall stage, and post radiation chemotherapy were included as covariates in a Cox Proportional Hazards Model for overall and cancer specific survival.

The multivariable Cox proportional hazards model for overall survival formed the basis of a nomogram to predict 1, 2, 5, and 8 year survival. The nomogram preserves the underlying relationship between the coefficients in the multivariable Cox proportional hazards model and merely maps these coefficients to easily interpretable point values. Predicted median survival was calculated as a weighted sum of the individual survival predictions. This model was validated using bootstrap resampling of the original patient cohort. This validation method creates new Cox models by drawing patients from the original cohort with replacement (bootstrap sample). The performance of bootstrap sample and original cohort against the bootstrap model are subtracted. This is repeated 1, 000 times, and all the differences are averaged. This value is subtracted from the performance of the original model against the original cohort creating an optimism corrected estimate. Alignment of the plotted optimism corrected estimate with the plotted performance of the model on the original cohort is an indication of internal validity and lack of overfitting (**Supplemental Figure 4**). Internal validation by bootstrap resampling has been shown to have stable performance estimates and low bias compared to other methods^13^.

To assess the relative contribution of each covariate towards prediction ability of the overall survival multivariable Cox model, the drop in C index was calculated by omitting each covariate one at a time from the full model. The C index has been routinely used to evaluate the discriminating ability of a survival model^14^.

### 2.3 Risk Groups

The aforementioned model development was the primary analysis of this study. Secondary analysis aimed to understand whether PHOM score may have potential to create risk groups. This cohort was divided into 2 groups by the median PHOM score of the entire cohort (PHOM = −0.0372). Kaplan-Meier curves were drawn to compare overall survival between the two groups, and log-rank statistics were used to compare the survival trends. Cumulative incidence curves were drawn to compare cause of death between these 2 groups.

This analysis was repeated with optimized tertiles. Tertiles were calculated by selecting the two PHOM score cutoff values that resulted in the maximal chi square value of the Kaplan-Meier Curve. The low risk group (n = 211) was defined as below -0.460, medium risk group (n = 172) as between -0.460 and 0.668, and high risk group (n = 171) as above 0.668. Both Kaplan Meier curves and cumulative incidence curves were drawn for these three groups. The Benjamini-Hochberg correction was used to correct for multiple comparisons for the Kaplan Meier pairwise comparisons. Post-hoc Kaplan Meier curves were drawn in a similar method to assess PHOM risk stratification in clinical subgroupings by KPS, CCI, and overall stage.

### 2.4 Data Availability

All data were processed and analyzed using R (v3.6.1) and Python (v3.7.6) ^15–17^. Anonymized data may be requested from the authors. Code with explanation for this data pipeline can be found at https://github.com/eashwarsoma/phom-lungca-ccf.

## 3 Results

### 3.1 Patient Characteristics

The patients in this study were generally able-bodied individuals with early stage NSCLC and a moderate number of comorbidities. **Table 1** provides a descriptive overview of the patient population. All patients had stage I or II primary NSCLC (none had nodal disease). No patients in this cohort had treatment prior to stereotactic body radiation therapy (SBRT), but 26 patients (4.7%) had some form of post SBRT chemotherapy. Karnofsky Performance Status (KPS) in this study ranged from 50 − 100. Most patients had baseline independent functional status corresponding to a KPS of 80 (215 patients) and 90 − 100 (168 patients). Using an *α/β* ratio of 10, mean dose received over treatment course was 100.54*Gy* (SD: 25.24*Gy*) EQD2. 89 patients had minimal comorbidity corresponding to a Charlson Comorbidity Index (CCI) of 2 − 4. Most patients had additional comordbities with scores ranging from 5 − 8. 60 patients had severe comorbidity (CCI *>* 8). 115 (20.8%) patients died from cancer, and 323 (58.3%) patients died from other causes. 116 patients (20.9%) were alive at last follow up, and the median follow up time for those alive was approximately 4.7 years (SD: 2.14 years).

**Table 1.**
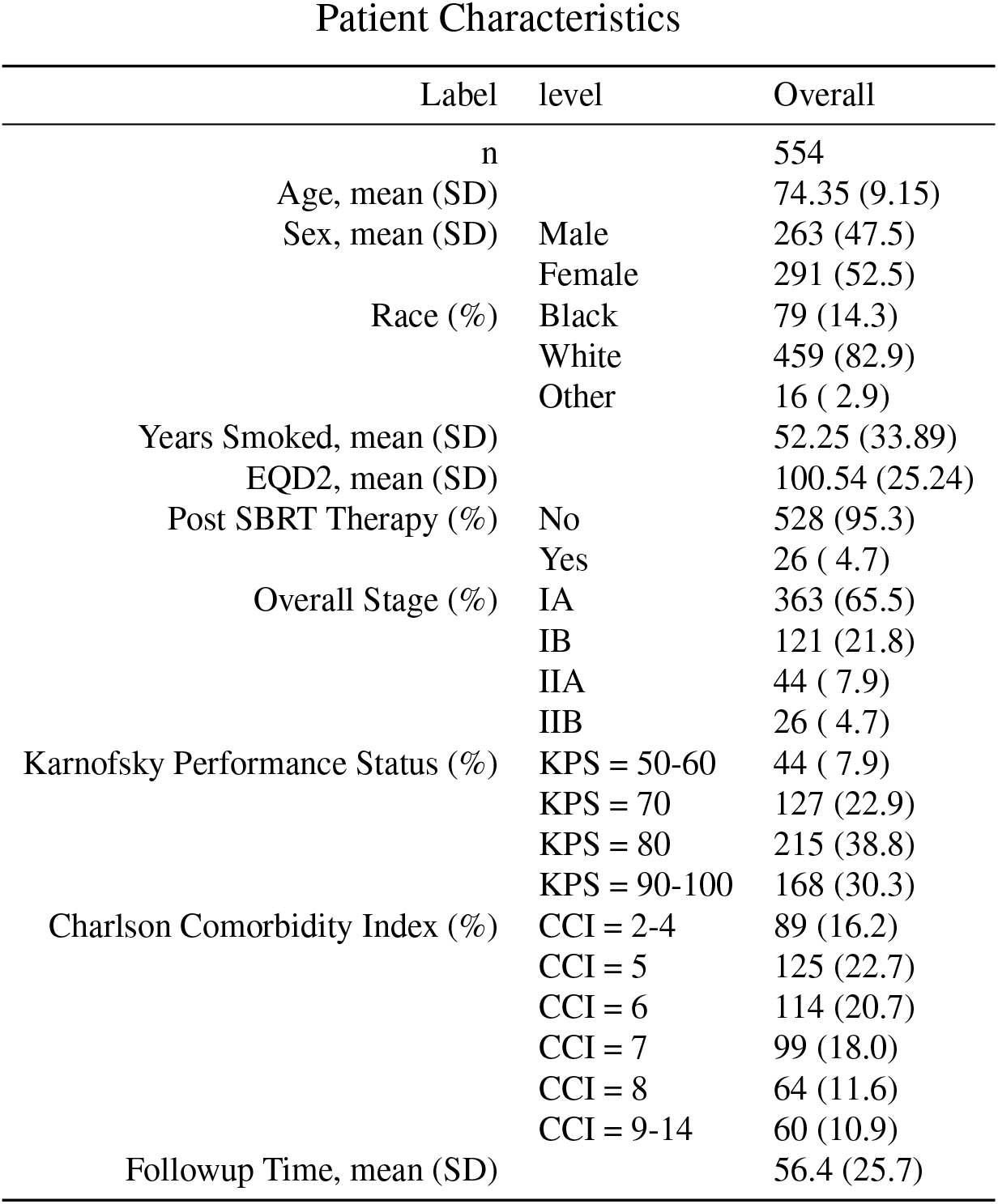
Patient Characteristics of Study Cohort. Patients in this cohort had early stage tumors, were generally healthy (high KPS), and had a moderate number of comorbidities (CCI: 5-8). 554 patients were present in the cohort. Followup time describes the mean number of months between treatment and most recent followup appointment for patients who were alive. EQD2: Equivalent Dose In 2-Gy Fractions, calculated using an *α/β* ratio of 10. KPS: Karnofsky Performance Score CCI: Charlson Comorbidity Index

### 3.2 Survival Models

PHOM score was a significant predictor of both overall survival (HR: 1.17; 95% CI: 1.07 − 1.28, *p <* 0.001) and cancer specific survival (HR: 1.31; 95% CI: 1.11 − 1.56, *p* = 0.0014) in their respective multivariable Cox models. **Table 2** lists the exact hazard ratios for all Cox models, and **Supplemental Figure 1** provides a forest plot visualization of the data. **Supplemental Table 1** provides associated p-values. For overall survival, KPS = 90 − 100 was associated with improved survival (HR: 0.46, 95% CI: 0.32 − 0.67, *p <* 0.001), and increased comorbidity was associated with poor overall survival (HR: 2.54, 95% CI: 1.69 − 3.80, *p <* 0.001).

**Table 2.**
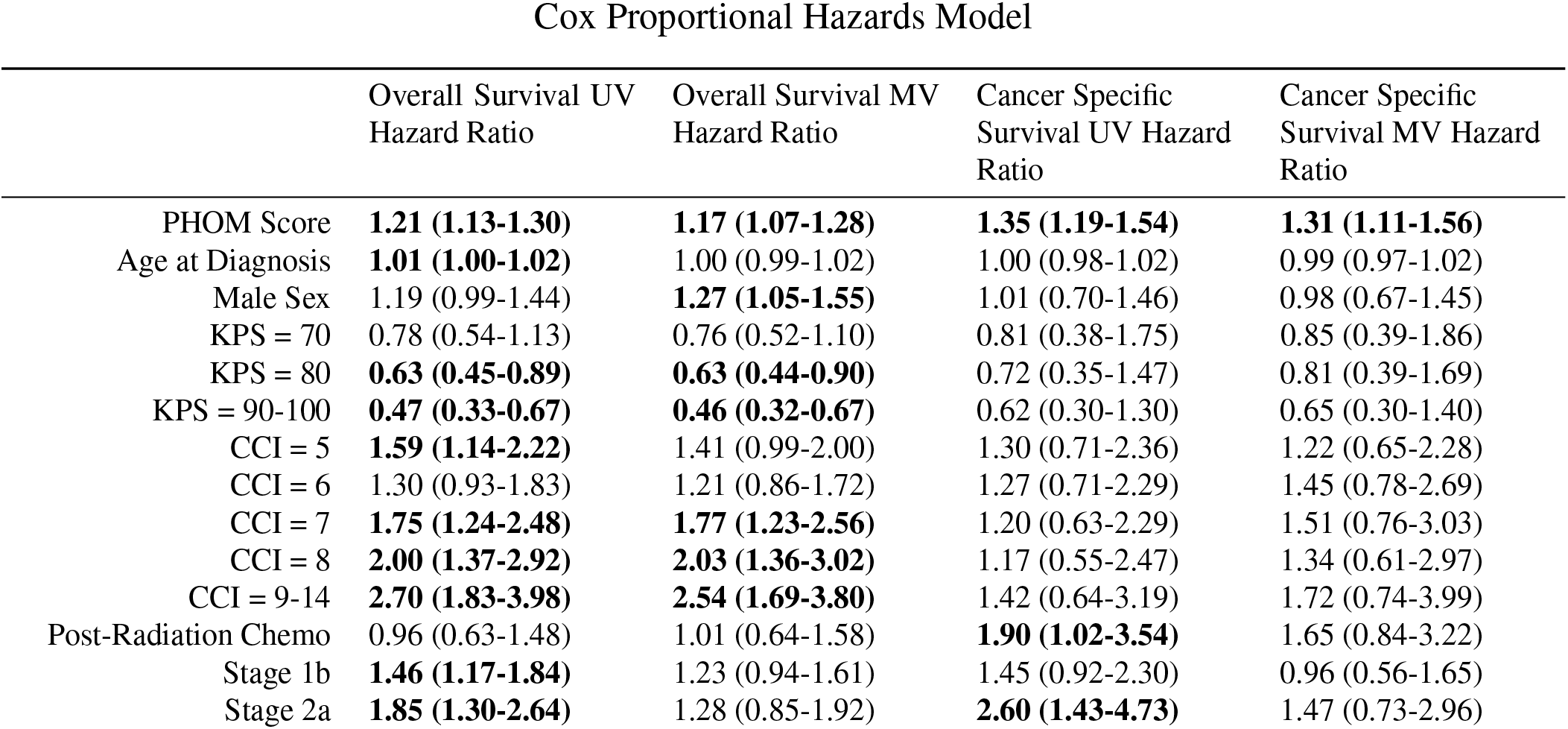
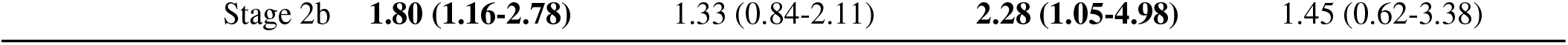
Cox Proportional Hazards Model for Overall and Cancer Specific Survival. The PHOM Score was a significant covariate for survival even after controlling for clinical variables. KPS and CCI were significant covariates for overall survival but not in cancer specific survival. Values are given as hazard ratio (95% Confidence Interval). KPS (Karnofsky Performance Status) is compared against KPS = 50 −60. CCI (Charlson Comorbidity Index) is compared against CCI of 2 −4. Stage covariates are compared against Stage 1a. Bolded values are statistically significant with *p <* 0.05. P-values for this table are included as **Supplemental Table 1**.

We drew nomograms, which are user friendly visual tools that represent the underlying survival regression models. Nomograms were calibrated for 1, 2, 5, and 8-year overall survival as shown in **Supplemental Figure 5**. The model was internally validated using bootstrap resampling, and calibration curves are shown in **Supplemental Figure 4**. The apparent and optimism bias corrected curves showed relatively strong alignment with the ideal curve indicating that our model is internally valid and not overfitted. **Supplemental Table 2** demonstrates the relative contribution of each covariate to the predictive ability of the multivariable Cox model with relative drop in C index. **Supplemental Figure 3** shows the proportion chi square contributed by each variable in the multivariable Cox Model. Both analyses revealed that PHOM Score, KPS, and CCI were the most important variables in the model and that each of these variables contributed unique information to the predictive ability of our model.

Each covariate from the multivariable Cox proportional hazards model is associated with a certain point value, and summing the point values for a given patient’s parameter generates a score that can be used to assign 1, 2, 5, and 8 year overall survival chance. The weighted sum of these survival chances gives the predicted median survival. An online interactive version of this nomogram is available at https://eashwarsoma.shinyapps.io/LungCancerTDATest/. We provide the exact risk point values associated with each clinical variable in this nomogram as a supplemental CSV file: Predictors_Points_Survival.csv.

### 3.3 Risk Stratification

The PHOM score produced well defined risk categories for overall survival and cancer death incidence when stratified by median and optimized cutoff values. **Figure 2A** shows patients with PHOM scores below the cohort median had a median survival of 45.4 months (95% CI: 40.1 − 51.8), which was significantly higher compared to patients in the high PHOM score group (median: 29.2 months, 95% CI: 23.6 − 34.3, *p <* 0.001). **Figure 2B** shows survival differences predicted by optimized PHOM score cutoffs. The median survival for the high PHOM risk group was 25.7 months (95% CI: 20.9 − 31.0). The median survival for the medium and low risk groups were 34.8 months (95% CI: 30.0 − 44.9) and 47.0 months (95% CI: 40.9 − 58.1) respectively. These groups had significant differences in overall survival (*p <* 0.001); however, the p-value is somewhat moot as the cutoffs were calculated by optimizing the chi square value of the Kaplan Meier curve. We applied these cutoffs to clinical groups stratified by KPS, CCI, or overall stage in **Supplemental Figure 2**. This post-hoc analysis demonstrated that higher PHOM risk score cutoffs were still associated with poorer survival in all clinical strata except for patients in Stage IB, IIA, and IIB where the trend was similar but did not reach statistical significance.

**Figure 2.**
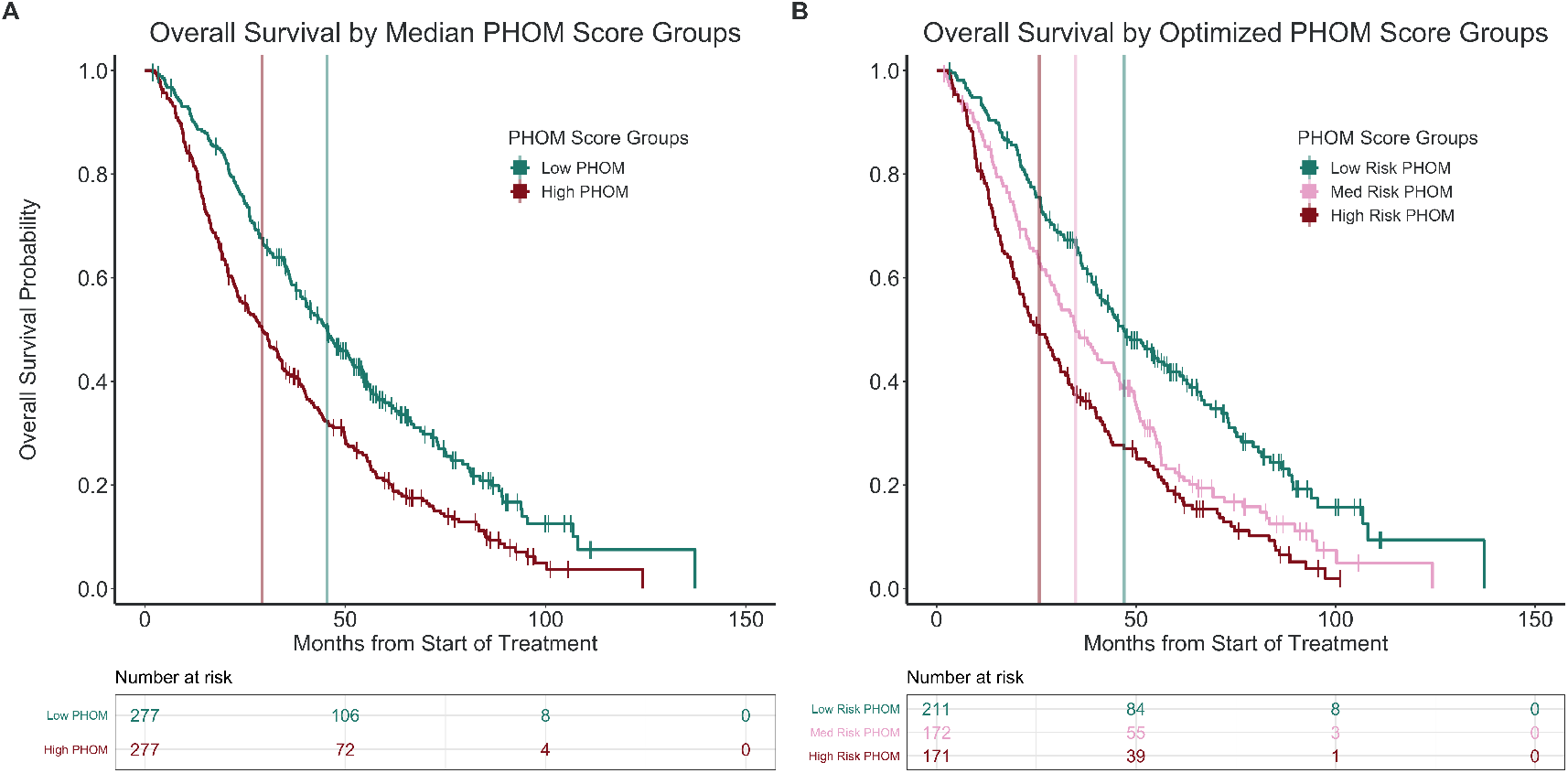
Kaplan Meier Overall Survival by PHOM Score. Stratifying the cohort by median PHOM score groups and optimized cutoffs produced well demarcated survival curves. **(A)** The median survival for the high PHOM group was 29.2 months (95% CI: 23.6 −34.3). The median survival for the low PHOM group was significantly higher at 45.4 months (95% CI: 40.1 −51.8, *p <* 0.001). **(B)** The median survival for the high PHOM risk group was 25.7 months (95% CI: 20.9 −31.0). This was significantly less than both the medium (*p* = 0.020) and low risk PHOM score groups (*p <* 0.001). The median survival for the medium risk group was 34.8 months (95% CI: 29.9 −44.9). This was significantly less than the low risk PHOM score group (*p <* 0.001). The median survival for the low risk group was 47.0 months (95% CI: 40.9 −58.1). However, it is important to note that these p-values are somewhat moot since the tertiles were calculated to maximize the chi square value of the KM curve.

**Figure 3A** compares cause specific death between PHOM score groups divided by cohort median. Having an above median PHOM score was associated with a 0.244 (95% CI: 0.192 − 0.296) chance of cancer specific death by month 65, which was significantly higher compared to the below median PHOM score chance of death of 0.171 (95% CI: 0.123 − 0.218, *p* = 0.029). However, for other causes of death, a high vs low PHOM score corresponded to a nonsignificant 0.582 (95% CI: 0.521 − 0.642) vs 0.499 (95% CI: 0.436 − 0.563) chance of death by month 65. **Figure 3B** compares cause specific death among PHOM score groups divided by optimized PHOM score risk groups. For low, medium, and high risk PHOM score groups, the chance of cancer death by month 65 was 0.146 (95% CI: 0.0950 − 0.197), 0.194 (95% CI: 0.133 − 0.255), and 0.296 (95% CI: 0.224 − 0.366) respectively (*p* = 0.0013). For low, medium, and high risk PHOM score groups, the chance of death from other causes by month 65 was 0.474 (95% CI: 0.401 − 0.546), 0.612 (95% CI: 0.535 − 0.689), and 0.551 (95% CI: 0.474 − 0.629) respectively.

**Figure 3.**
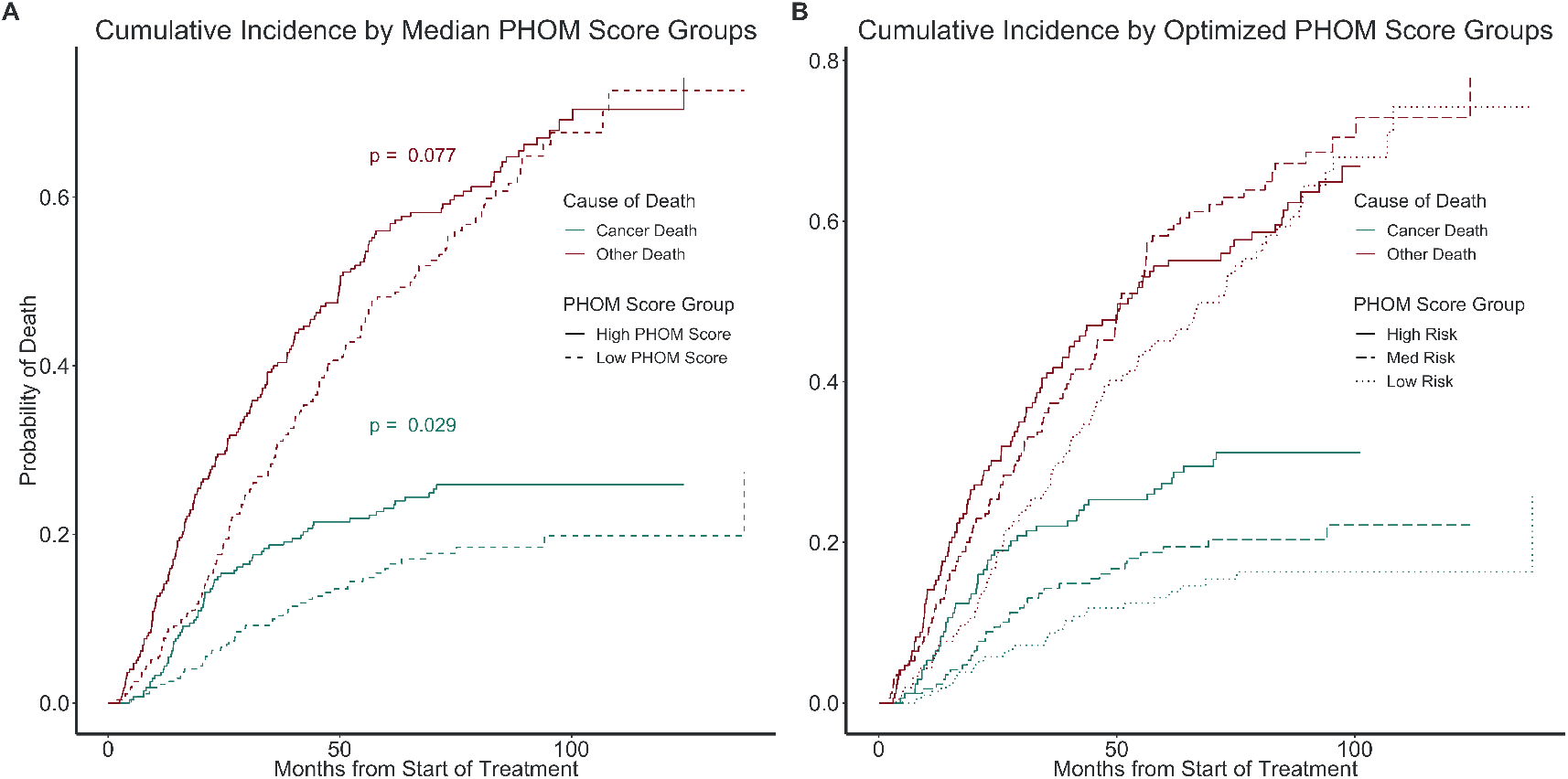
Cumulative Incidence of Death by Specific Cause. Stratifying the cohort by median PHOM score groups and optimized cutoffs produced well demarcated cumulative incidence curves for cancer specific death but not death due to other causes. **(A)** Having an above median PHOM score was associated with a 0.244 (95% CI: 0.192 −0.296) chance of cancer specific death by month 65. A below median PHOM score was associated with a significantly less chance of death by month 65 at 0.171 (95% CI: 0.123 −0.218, *p* = 0.029). However, for other causes of death, a high vs low PHOM score corresponds to a nonsignificant 0.582 (95% CI: 0.521 −0.642) vs 0.499 (95% CI: 0.436 −0.563) chance of death by month 65. **(B)** For low, medium, and high risk PHOM score groups, the chance of cancer death by month 65 was 0.146 (95% CI: 0.0950 −0.197), 0.194 (95% CI: 0.133 −0.255), and 0.296 (95% CI: 0.224 −0.366) respectively. For low, medium, and high risk PHOM score groups, the chance of death from other causes by month 65 was 0.474 (95% CI: 0.401 −0.546), 0.612 (95% CI: 0.535 −0.689), and 0.551 (95% CI: 0.474 −0.629) respectively. Similarly, the difference among incidence of death at month 65 was significant when assessing cancer death (*p* = 0.0013) but not other death. 115 patients in this cohort died of cancer specific death of which 80 were due to nodal and distal progression and the remaining 35 due to new primary, local, or lobar progression. The p-values are moot similar to the Kaplan-Meier risk stratification analysis since we maximized over the chi square value of the KM curve.

## 4 Discussion

### 4.1 Strengths of the PHOM Score

Our novel topologically inspired radiomics variable predicts survival and risk stratifies patients with NSCLC. To our knowledge, there is currently no similar topology based imaging metric that has demonstrated survival prediction in patients with NSCLC.

Stratifying PHOM score by the cohort median reveals worse overall survival and cancer specific survival in the high PHOM score group. PHOM score was also the only significant predictor in the multivariable cancer specific Cox model. Additionally, it differentiated cancer-specific death from death by other causes suggesting that the PHOM score is truly measuring cancer severity (**Figure 2** and **Figure 3**).

Interestingly, our post-hoc analysis in **Supplemental Figure 2** revealed that PHOM score continued to stratify survival across all clinical subgroupings of KPS and CCI. This is expected as PHOM score is a measure of the tumor itself whereas KPS and CCI are measures of overall health. Stage did not contribute to predicting survival in the multivariable models, possibly due to high local control rates when early stage tumors are treated with SBRT^18^. However, in **Supplemental Figure 2G**, the PHOM score identified high risk patients within the Stage IA group. While Stage IA patients are thought to be low risk overall, approximately 30% of patients with this stage do not survive over a five year interval^19^. We need better risk stratification, and we offer the PHOM score tertiles as a possibility. Risk stratifying early stage NSCLC cancers can signal a need for adjuvant systemic therapy and assist clinical trialists in creating treatment groups based on risk. However, further study is needed as this was a post-hoc analysis. While stages IB, IIA, and IIB had similar trends when grouping by PHOM risk scores, it did not reach statistical significance.

The PHOM score’s greatest strength is its ability to predict overall survival. The C-index demonstrates the predictive ability of a model. When omitting variables one at a time from the model, we found that KPS had the greatest drop in C-index score, and CCI and PHOM score resulted in similar C-index drops. This reflects the importance and unique information each covariate brings to the model (**Supplemental Table 2**). The calibration plot show demonstrates strong internal validity of our nomogram’s predictive ability (**Supplemental Figure 4**).

### 4.2 Clinical Implications

To our knowledge, this is the first paper that utilizes persistent homology to predict overall survival in NSCLC. Our prior work was limited to demonstrating statistical association with overall survival and did not include important covariates such as KPS and CCI due to limitations in available data^12^. We speculate our metric measures underlying structural tissue heterogeneity. Increased tissue heterogeneity may indicate aggressive tumor growth providing an explanation of why our metric correlates well with cancer specific survival and predicts overall survival. Direct pathology data and tumor marker data were not available in this study but should be assessed in future studies to define the biological significance of the PHOM score. Our new imaging persistent homology metric adds to the other metrics that have been developed for glioblastoma, breast, and prostate cancer clinical outcomes research^8,10,20^.

Persistent homology has opened up a new suite of tools for image analysis. We have shown that the PHOM score predicts survival in NSCLC even after controlling for clinical metrics. How the PHOM score could interact with traditional radiomic measures remains to be answered. Additionally, the PHOM score could be incorporated with non-imaging data. Genomic models such as the genomically adjusted radiation dose (GARD) predict individualized response to different radiation doses as demonstrated in a meta-analysis^21,22^. We foresee future clinical models that incorporate genomic, imaging, and other data modalities to more precisely guide therapy.

### 4.3 Limitations

While our results are promising for NSCLC, some limitations must be considered. Our nomogram is only validated for patients with early stage NSCLC who receive SBRT therapy as primary treatment. While we do not see a reason for this metric to not work for surgical patients, we cannot formally say our nomogram is validated for this population. The patients in this cohort all received care at a tertiary healthcare center and sometimes became candidates for SBRT due to comorbidities contraindicating surgery. As such, they are more likely to be sicker than the average patient with early stage NSCLC, which is reflected in our overall survival rates being less than typical median survival of over 5 years^23^. CT standards vary among institution, so it is not clear whether sources of imaging variance have an impact on PHOM score. However, our prior work assessed scans with varied parameters different from this institution’s parameters and was still successful suggesting a potential resilience of this score to this particular confounder^12^.

We could not generate a cancer specific survival curve due to poor calibration curves. We believe this to be a result of the limited number of cancer specific deaths–likely due to successful treatment of early stage tumors. For completeness, we include these attempted curves in **Supplemental Figure 6**.

### 4.4 Conclusion

Publicizing this nomogram, recruiting multi-institutional data, and conducting prospective studies could address these limitations in future studies. Additionally, future work could analyze how the PHOM score interacts with traditional radiomics. The widespread imaging capability of modern hospitals makes imaging metrics such as the PHOM score practical to utilize in patient care. In cohort studies or randomized controlled trials, the PHOM score could be used to risk stratify patients to investigate who might benefit from adjuvant therapy. Long term, we envision the creation of a risk calculator that extracts the most useful features of imaging data to both prognosticate and guide clinical decisions.

## Data Availability

All data produced in the present study are available upon reasonable request to the authors.

https://github.com/eashwarsoma/phom-lungca-ccf

https://eashwarsoma.shinyapps.io/LungCancerTDATest/

## Conflicts of Interest

The authors have no conflicts of interest to disclose, financial or otherwise, in relation this article.

## Funding

JGS was supported by NIH grant R37 CA244613 and American Cancer Society. ES and RRW were supported by the Case Comprehensive Cancer Center Summer Training Grant for medical students. KY was supported by the Computational Genomic Epidemiology of Cancer (CoGEC) Program at Case Comprehensive Cancer Center (T32CA094186), Young Investigator Award from ASCO Conquer Cancer Foundation, and RSNA Research Resident Grant.

## Acknowledgments

The authors thank members of Theory Division for their constructive feedback and valuable input in this project. We thank Jessica Scarborough, PhD for advice and code to generate optimal PHOM score tertiles. JGS was supported by NIH grant R37 CA244613, NIH grant U54 (Radiation Oncology Biology Integration Network), and the American Cancer Society through their Research Scholar Grant. ES and RRW were supported by the Case Comprehensive Cancer Center Summer Training Grant for medical students. KY was supported by the Computational Genomic Epidemiology of Cancer (CoGEC) Program at Case Comprehensive Cancer Center (T32CA094186), Young Investigator Award from ASCO Conquer Cancer Foundation, and RSNA Research Resident Grant.

## Author contributions statement

All authors made substantial contributions to: (1) the conception and design of the study or acquisition, analysis, and interpretation of the data; (2) draft the article or revising it critically for important intellectual content; (3) final approval of the submitted version.

## Supplemental Information

**Supplemental Figure 1** shows the forest plot and **Supplemental Table 1** lists the exact values for the hazard ratios across all Cox proportional hazards models. The PHOM score was the only covariate that was significant in all four models suggesting it is a predictor of overall and cancer survival.

**Supplemental Figure 1.**
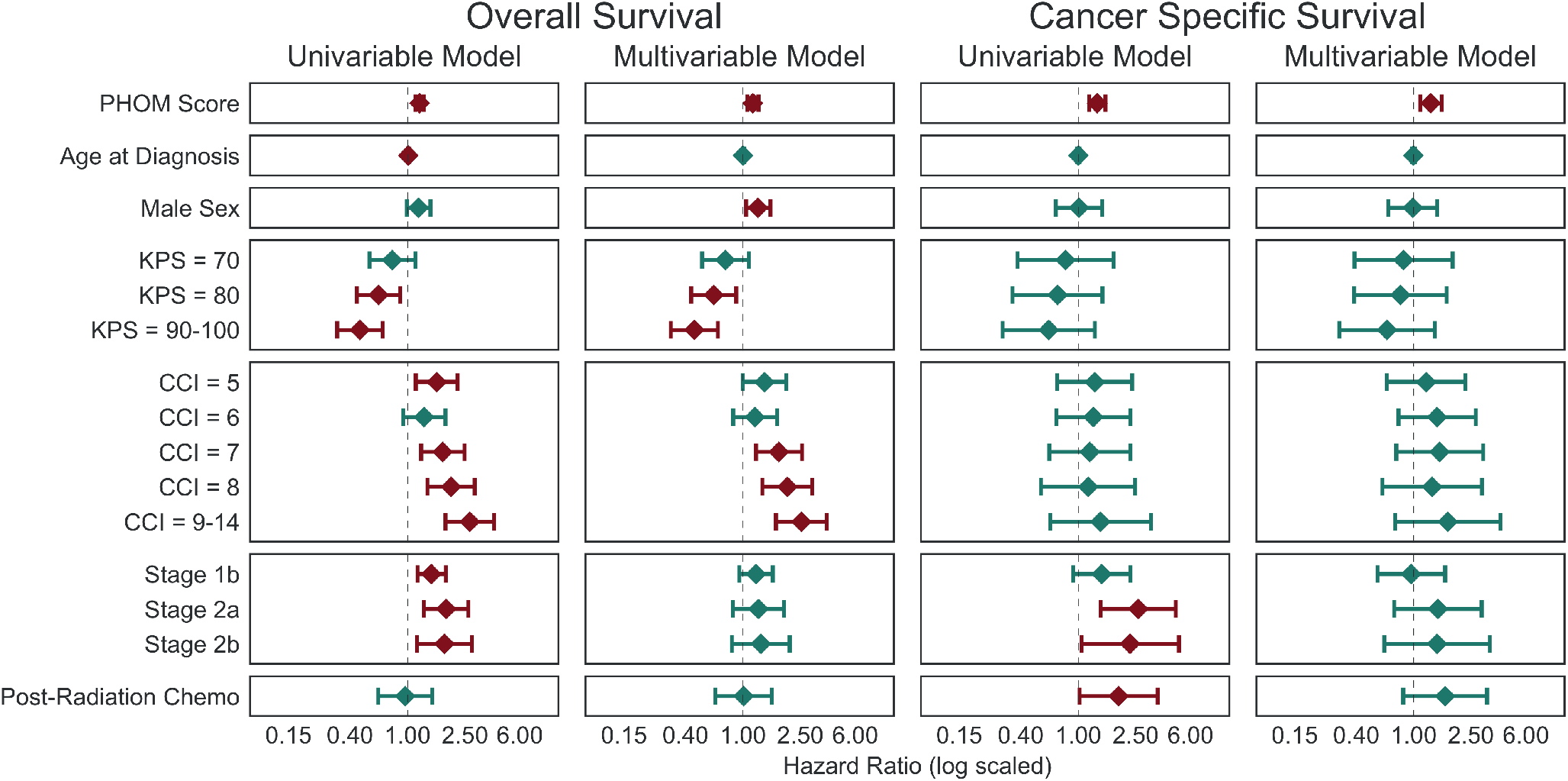
Forest Plot of Overall and Cancer Specific Cox Proportional Hazards Models. The PHOM Score was the only significant covariate for both overall and cancer specific survival. Bands represent 95% confidence intervals. The red bars represent significant hazard ratios. KPS (Karnofsky performance status) is compared against KPS = 50 −60. CCI (Charlson Comorbidity Index) is compared against CCI = 2 −4. Stage covariates are compared against Stage 1a.

**Supplemental Table 1.** Cox Proportional Hazards Model for Overall and Cancer Specific Survival with p-values. Values given as Hazard ratio (95% Confidence Interval). KPS (Karnofsky performance status) is compared against KPS *>* 70. CCI (Charlson Comorbidity Index) is compared against CCI of 1-4. Stage covariates are compared against Stage 1a. This is the same table as P-values for this table are included as **Table 2** with p-values added.

|  | Overall Survival Univariable Model |  | Overall Survival Multivariable Model |  | Cancer Specific Survival Univariable Model |  | Cancer Specific Survival Multivariable Model |  |
| --- | --- | --- | --- | --- | --- | --- | --- | --- |
|  | Hazard Ratio | p-value | Hazard Ratio | p-value | Hazard Ratio | p-value | Hazard Ratio | p-value |
| PHOM Score | 1.21<br>(1.13-1.30) | < 0.001 | 1.17<br>(1.07-1.28) | < 0.001 | 1.35<br>(1.19-1.54) | < 0.001 | 1.31<br>(1.11-1.56) | 0.0014 |
| Age at Diagnosis | 1.01<br>(1.00-1.02) | 0.0077 | 1.00<br>(0.99-1.02) | 0.46 | 1.00<br>(0.98-1.02) | 0.96 | 0.99<br>(0.97-1.02) | 0.61 |
| Male Sex | 1.19<br>(0.99-1.44) | 0.067 | 1.27<br>(1.05-1.55) | 0.017 | 1.01<br>(0.70-1.46) | 0.95 | 0.98<br>(0.67-1.45) | 0.94 |
| KPS = 70 | 0.78<br>(0.54-1.13) | 0.18 | 0.76<br>(0.52-1.10) | 0.14 | 0.81<br>(0.38-1.75) | 0.6 | 0.85<br>(0.39-1.86) | 0.68 |
| KPS = 80 | 0.63<br>(0.45-0.89) | 0.0087 | 0.63<br>(0.44-0.90) | 0.011 | 0.72<br>(0.35-1.47) | 0.37 | 0.81<br>(0.39-1.69) | 0.57 |
| KPS = 90-100 | 0.47<br>(0.33-0.67) | < 0.001 | 0.46<br>(0.32-0.67) | < 0.001 | 0.62<br>(0.30-1.30) | 0.21 | 0.65<br>(0.30-1.40) | 0.27 |
| CCI = 5 | 1.59<br>(1.14-2.22) | 0.0069 | 1.41<br>(0.99-2.00) | 0.055 | 1.30<br>(0.71-2.36) | 0.4 | 1.22<br>(0.65-2.28) | 0.54 |
| CCI = 6 | 1.30<br>(0.93-1.83) | 0.12 | 1.21<br>(0.86-1.72) | 0.28 | 1.27<br>(0.71-2.29) | 0.42 | 1.45<br>(0.78-2.69) | 0.24 |
| CCI = 7 | 1.75<br>(1.24-2.48) | 0.0015 | 1.77<br>(1.23-2.56) | 0.0022 | 1.20<br>(0.63-2.29) | 0.58 | 1.51<br>(0.76-3.03) | 0.24 |
| CCI = 8 | 2.00<br>(1.37-2.92) | < 0.001 | 2.03<br>(1.36-3.02) | < 0.001 | 1.17<br>(0.55-2.47) | 0.69 | 1.34<br>(0.61-2.97) | 0.47 |
| CCI = 9-14 | 2.70<br>(1.83-3.98) | < 0.001 | 2.54<br>(1.69-3.80) | < 0.001 | 1.42<br>(0.64-3.19) | 0.39 | 1.72<br>(0.74-3.99) | 0.21 |
| Had Post-Radiation Chemo | 0.96<br>(0.63-1.48) | 0.86 | 1.01<br>(0.64-1.58) | 0.97 | 1.90<br>(1.02-3.54) | 0.043 | 1.65<br>(0.84-3.22) | 0.14 |
| Stage 1b | 1.46<br>(1.17-1.84) | < 0.001 | 1.23<br>(0.94-1.61) | 0.12 | 1.45<br>(0.92-2.30) | 0.11 | 0.96<br>(0.56-1.65) | 0.88 |
| Stage 2a | 1.85<br>(1.30-2.64) | < 0.001 | 1.28<br>(0.85-1.92) | 0.24 | 2.60<br>(1.43-4.73) | 0.0017 | 1.47<br>(0.73-2.96) | 0.28 |
| Stage 2b | 1.80<br>(1.16-2.78) | 0.0082 | 1.33<br>(0.84-2.11) | 0.23 | 2.28<br>(1.05-4.98) | 0.038 | 1.45<br>(0.62-3.38) | 0.39 |

**Supplemental Figure 2** utilizes the PHOM score risk cutoffs calculated from the entire cohort (see **Figure 2** to assess risk stratification within clinical subcohorts. The PHOM Score continues to risk stratify across all KPS and CCI groups, which is expected as the former is a measure of tumor property and the latter are measures of overall health. Interestingly, PHOM stratifies risk by stage (another tumor property) for IA but not others. This suggests that PHOM is able to identify high risk patients in the overall low risk Stage IA patients. However, this post hoc analysis is limited by multiple comparisons (though within each KM curve, pairwise comparisons are BH corrected) and limited sample size. In **Supplemental Figure 2H**, the relative order of risk groupings are the same suggesting PHOM score may also risk stratify this cohort but simply did not reach statistical significance. Additional data and research are needed to truly verify whether PHOM can risk stratify within clinical stages.

**Supplemental Figure 3** shows a graphical representation of the proportion chi-square each variable contributes to the final model. This metric assess the relative strength of association to survival outcome for each variable in the Cox proportional hazards model. **Supplemental Table 2** shows the drop in C-index in the rightmost column when a variable is omitted from the full model. For example, in the first row showing KPS, the C-index drop reflects the C-index of the full model minus the C-index of the model with all variables except KPS. Larger positive numbers drops indicate that particular variable contributes relatively more to the predictive ability of the model. Importantly, omitting one covariate at a time also tells us whether that covariate provides redundant predictive information. Since PHOM Score, KPS, and CCI resulted in the largest drops in C-index, these variables give the most predictive information in the multivariable Cox-proportional hazards survival model.

We internally validated the Cox models using bootstrap resampling. The apparent performance reflects the original model performance on boot strap samples. The bias-corrected performance represents the performance of the same multivariable Cox model but recalculated using a boot strap sample. The primary function of the bias-corrected line is to show our model’s prediction is not the result of overfitting on our data. Relative overlap between the apparent and bias-corrected lines along the ideal line (the hypothetical perfect model) represents strong internal validity of our model.

**Supplemental Table 2.**
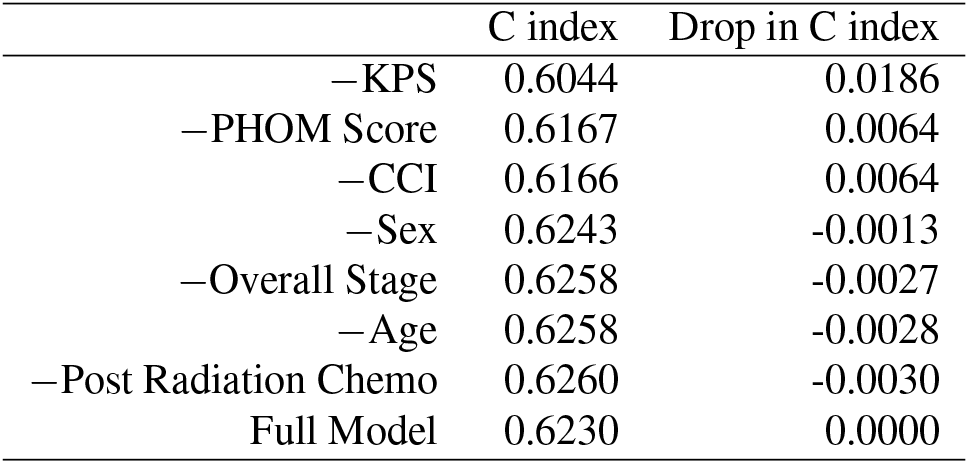
Predictive Ability of Each Variable for Overall Survival Cox Model. The bottom row describes the full model, and the other rows describe the models generated from knocking out the listed variable. C-index is a measure of assessing whether the risk predicted by the model is associated with reduced survival^14^. Drop in C-index was calculating by C-index of the full model − C-index of the one variable knockout models.

For completeness, we included the calibration curves we attempted for cancer specific death **Supplemental Figure 6**. Given the relative lower number of cancer deaths compared to deaths from other causes, the calibration curves had wide AUC (equivalent to C-index) confidence intervals. We opted not to develop a nomogram since these we felt the calibration curves were inadequate especially compared to the calibration curves for overall survival in **Supplemental Figure 4**.

**Supplemental Figure 2.**
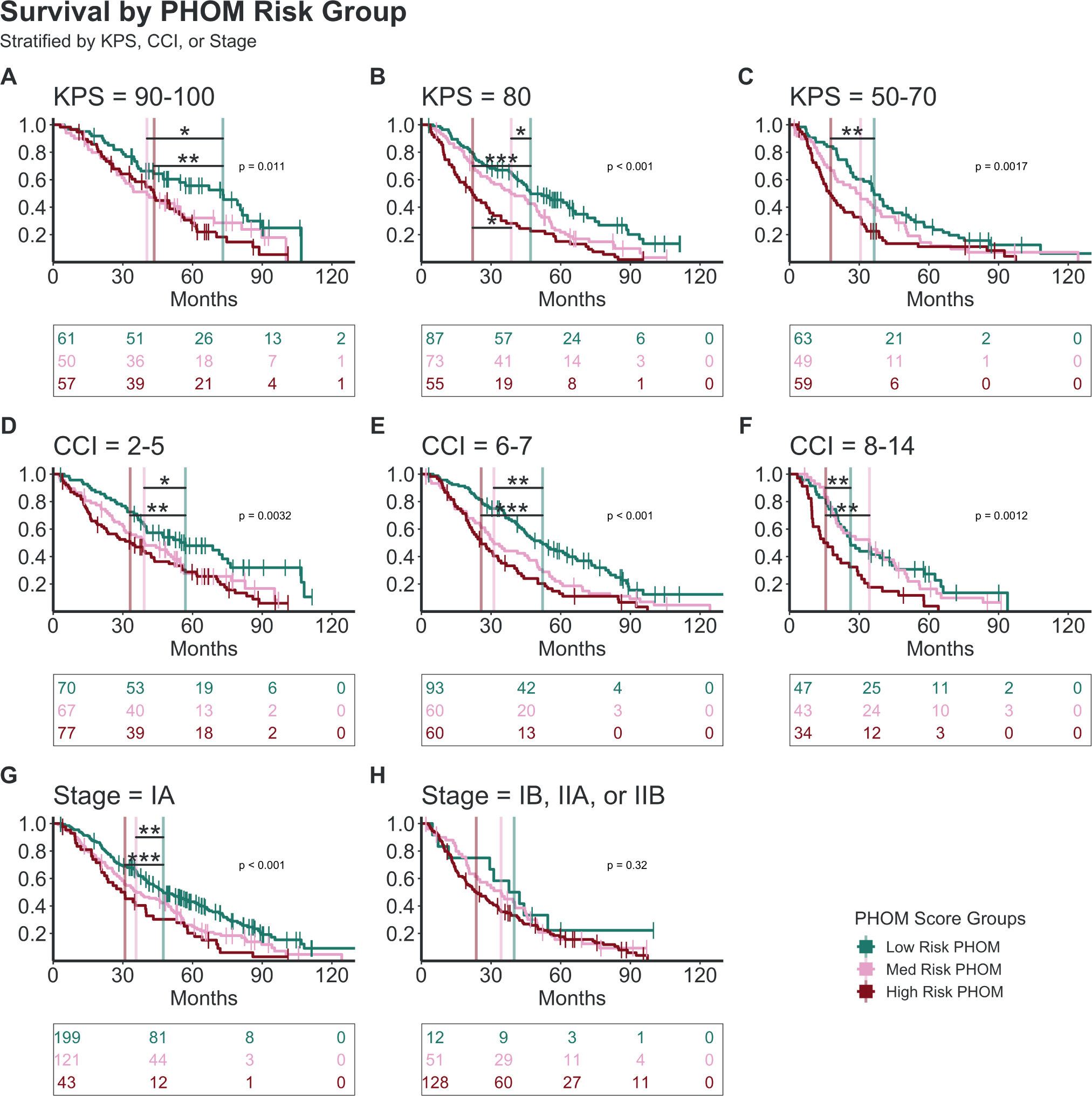
PHOM Risk Groups Stratified by Clinical Variables. The optimized PHOM risk cutoffs generated from the full cohort continue to risk stratify for patients subsetted by KPS (Karnofsky performance status) and CCI (Charlson Comorbidity Index) values. The PHOM score stratifies Stage IA patients but does not stratify patients in higher stages at a level of statistical significance. Pairwise comparisons between PHOM risk groups were Benjamini-Hochberg adjusted. Tables below each KM Curve reflect number of patients at risk at time points relative to the overlying x-axis.

**Supplemental Figure 3.**
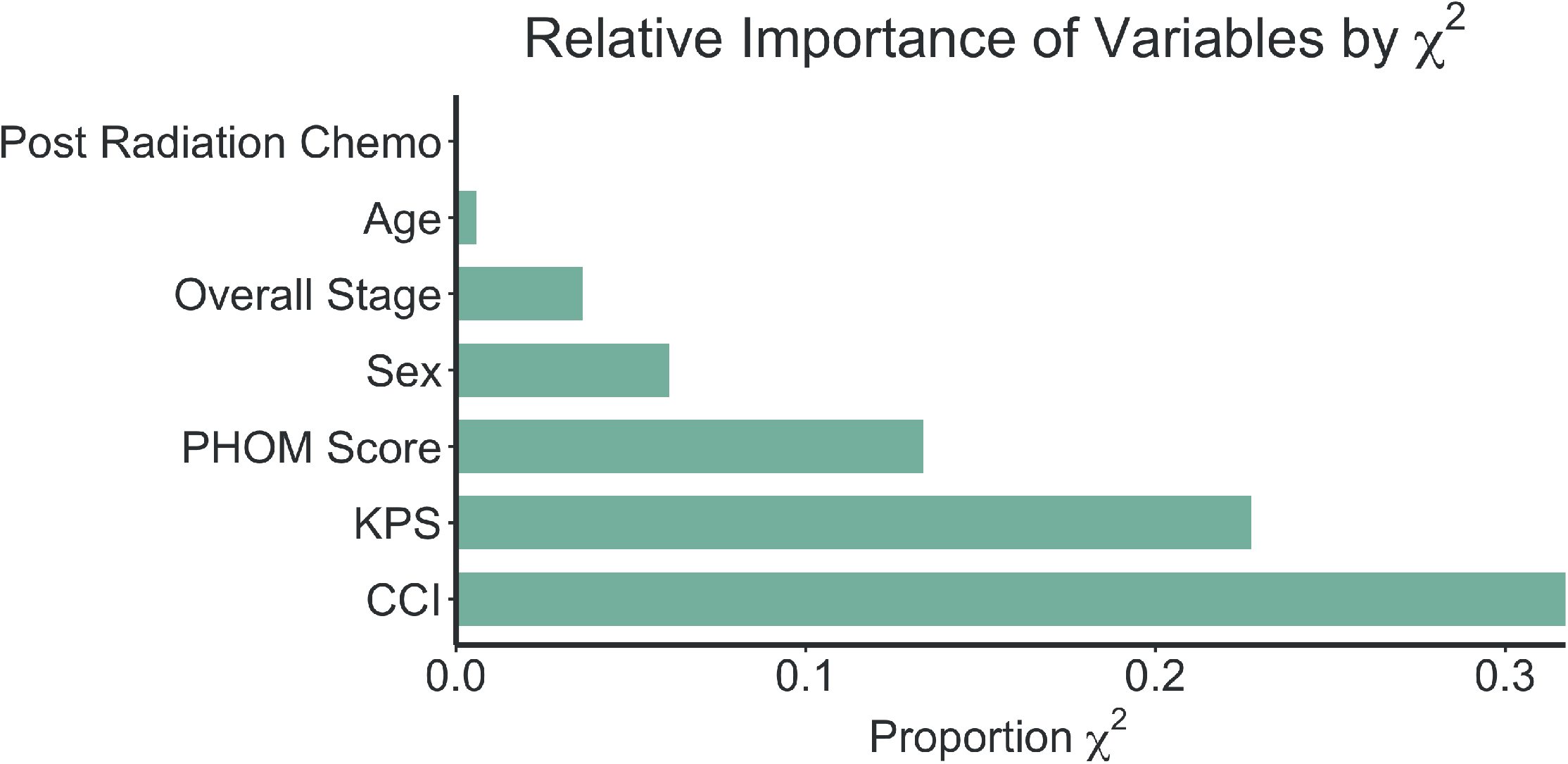
Proportion Chi Square of Each Predictor in Multivariable Cox Model. Proportion Chi Square reflects importance of variable in survival prediction of the model. KPS (Karnofsky Performance Status), CCI (Charlson Comorbidity Index), and PHOM Score were by far the most important predictors.

**Supplemental Figure 4.**
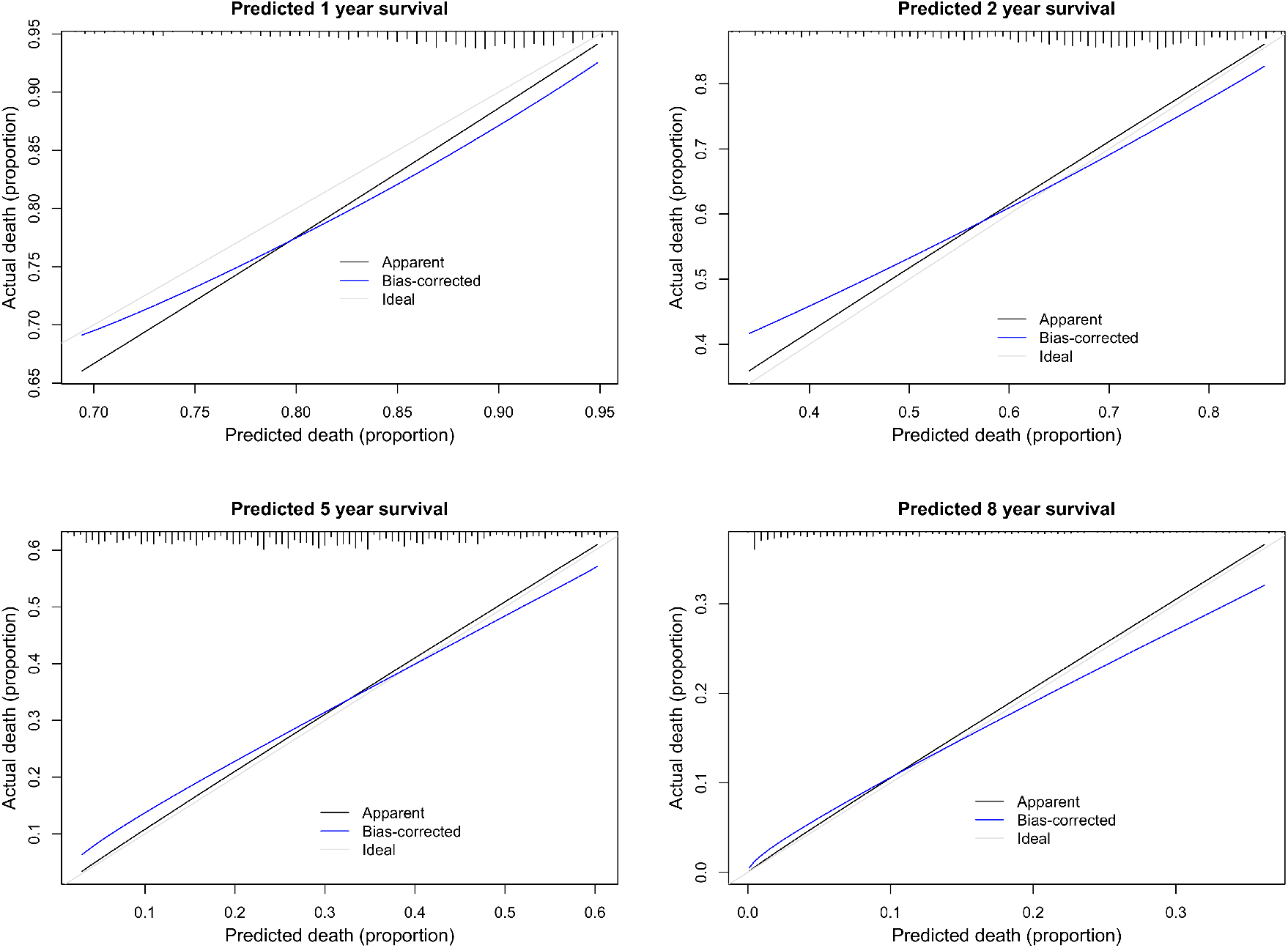
Calibration Curves for Overall Survival Cox Proportional Hazards Model. The apparent calibration line represents represents the performance of our model on the original data set. Bias corrected curves were calculated through 1000 bootstrap resamples. For each bootstrap sample, a new Cox model using the bootstrap sample was generated. Optimism was calculated as the difference between the performance of the real sample on the bootstrap Cox model and the performance of the bootstrap sample on the bootstrap Cox model. The optimism estimate from each bootstrap resample were averaged and subtracted from the apparent performance curve to generate the bias corrected curve. This bias corrected curve aligning with the apparent and ideal curves is an indication that our model is not overfitted.

**Supplemental Figure 5.**
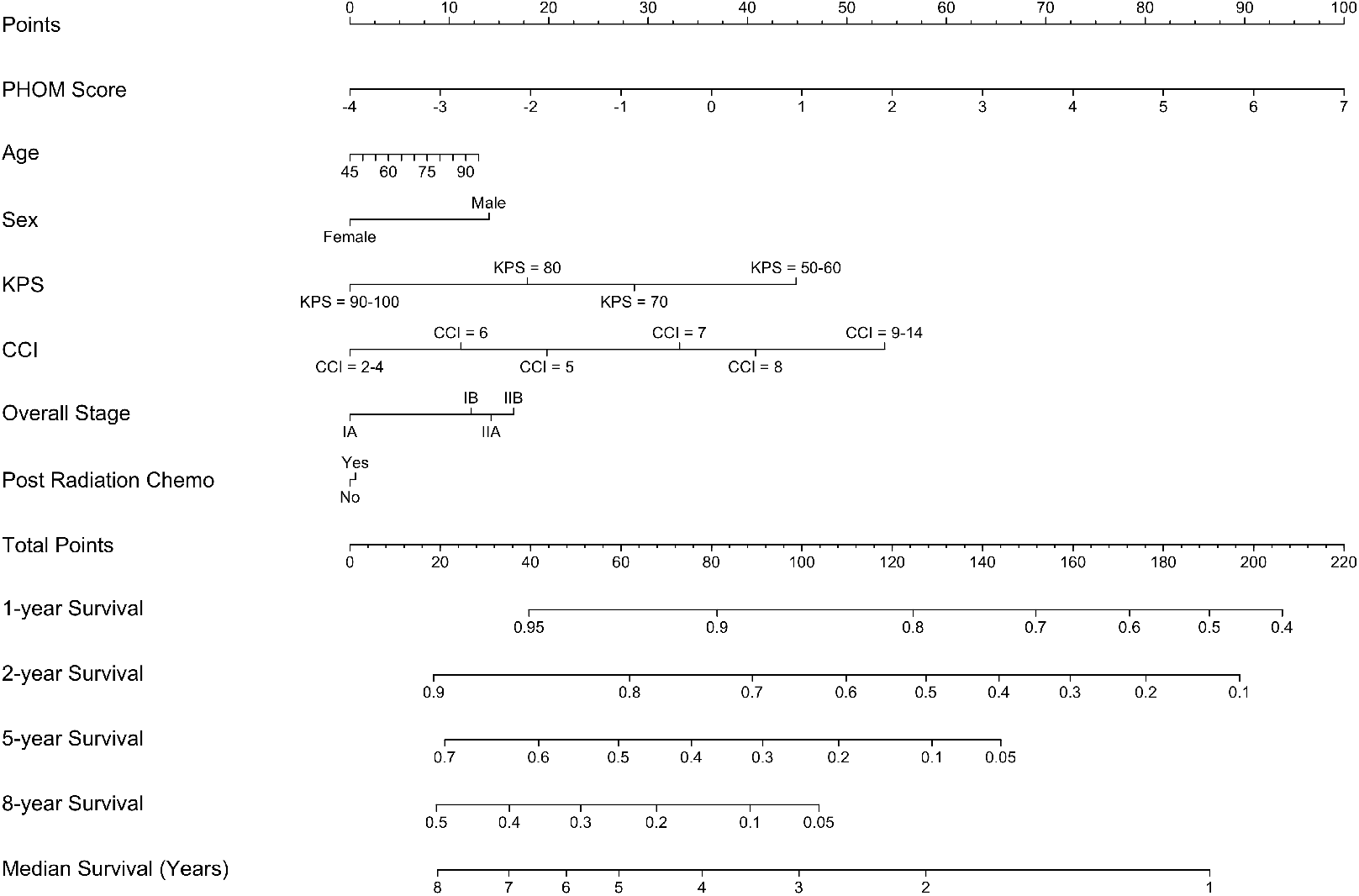
Nomogram for Overall Survival. This nomogram is a visual tool to convert the variables in our Cox overall survival model into a common risk score. This risk score can then convert into predicted survival. An online interactive version of this nomogram is available at https://eashwarsoma.shinyapps.io/LungCancerTDATest/. KPS: Karnofsky Performance Status. CCI: Charlson Comorbidity Index.

**Supplemental Figure 6.**
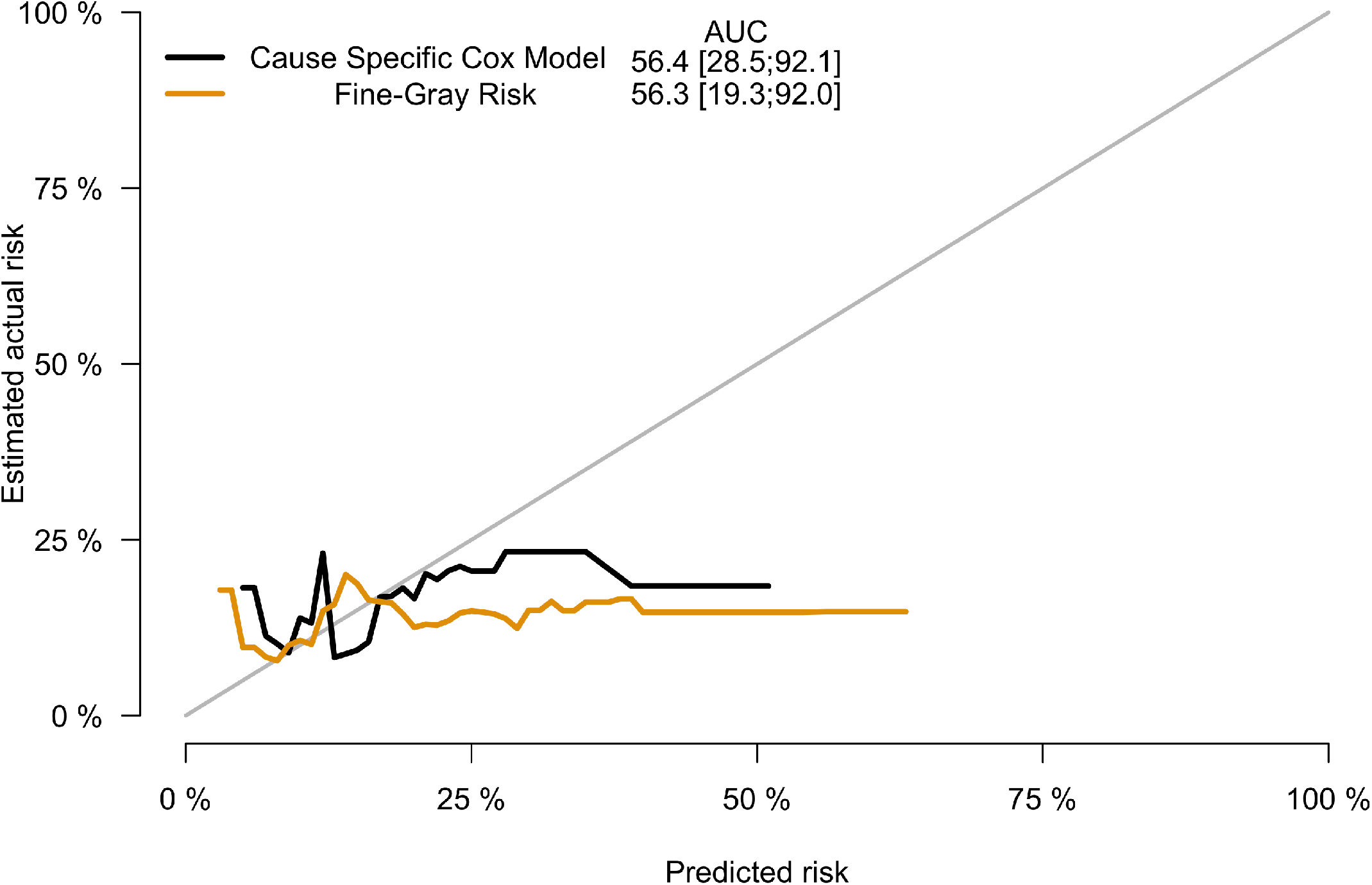
Calibration Curves for Cancer Specific Death Models. Both cause specific Cox and Fine-Gray Risk models did not produce adequate calibration curves so nomograms for cancer specific survival were not developed. AUC reported is equivalent to the C-index.

## Notes

### Competing Interest Statement

The authors have declared no competing interest.

### Author Declarations

IRB of Cleveland Clinic Foundation gave ethical approval for this work.

### Summary of Updates

-Modified title, clarifications in methodology, new supplemental figures and excel file

